# Environmental sampling for severe acute respiratory syndrome coronavirus 2 (SARS-CoV-2) during a coronavirus disease (COVID-19) outbreak aboard a commercial cruise ship

**DOI:** 10.1101/2020.05.02.20088567

**Authors:** Motoi Suzuki, Hajime Kamiya, Kiyoko Okamoto, Takuya Yamagishi, Kensaku Kakimoto, Makoto Takeda, Syutoku Matsuyama, Kazuya Shirato, Naganori Nao, Hideki Hasegawa, Tsutomu Kageyama, Ikuyo Takayama, Shinji Saito, Takaji Wakita, Makoto Ohnishi, Koji Wada, Retsu Fujita, Yoshiaki Gu, Nobuaki Matsunaga, Mikiyo Sakaguchi, R.N. Taichi Tajima, Norio Ohmagari, Saho Takaya, Hiroki Saito, Keiji Okinaka, Mathew Griffith, Amy Elizabeth Parry, Mateusz M Plucinski, Brenda Barnetson, James Leonard

## Abstract

**Background:** A COVID-19 outbreak occurred in a cruise ship with 3711 passengers and crew in 2020. This study is to test the hypothesis that environmental surfaces played important roles in transmission for SARS-CoV-2 during this outbreak.

**Methods:** We sampled environmental surfaces including air from common areas in the cruise ship and cabins in which confirmed COVID-19 cases and non-cases had stayed after they left the cabins. We tested the samples for SARS-CoV-2 by rt-PCR and conducted viral isolation.

**Findings:** Of 601 samples tested, SARS-CoV-2 RNA was detected from 58 samples (10%) from case-cabins from which they left 1-17 days before sampling, but not from non-case-cabins. Except for one sample from an air hood in a corridor, SARS-CoV-2 RNA was not detected from samples in common areas. SARS-CoV-2 RNA was not detected from all 14 air samples. RNA was most often detected on the floor around toilet in the bathroom (39%, 13/33, cycle quantification (Cq): 26.21-37.62) and bed pillow (34%, 11/32, Cq: 34.61-38.99). There was no difference in the detection proportion between cabins for symptomatic (15%, 28/189, Cq: 29.79-38.86) and asymptomatic cases (21%, 28/131, Cq: 26.21-38.99). No SARS-CoV-2 virus was isolated from any of the samples.

**Interpretation:** The environment around the COVID-19 cases was extensively contaminated from SARS-CoV-2 during COVID-19 outbreak in the cruise ship. Transmission risk of SARS-CoV-2 from symptomatic and asymptomatic patients seems to be similar and the environmental surface could involve viral transmission through direct contact.

## Background

Transmission of infectious disease aboard cruise ships is not a new issue. Easily transmittable viruses such as norovirus, have a long relationship with cruise ship outbreaks^1^. One possibility for this relationship is its mass-gathering characteristics that many people have a chance of close contacts and the other is contribution of environment to transmission. Environmental analysis of cruise ships found sanitary conditions aboard the inspected ships was often inadequate^2^.

On 2 February 2020, Hong Kong health authorities notified Japanese health authorities through the International Health Regulation mechanism that a passenger who had been aboard a commercial cruise ship had disembarked in Hong Kong on 25 January and tested positive for severe acute respiratory syndrome coronavirus 2 (SARS-CoV-2)^3^. The vessel, which had 2666 passengers and 1045 crew aboard, was arriving at Yokohama, Japan, and on 3 February Japanese authorities ordered all passengers and crew to remain aboard. On 3 and 4 February, health officials obtained oropharyngeal specimens from those who had a fever or respiratory symptoms^4^. On 5 February, 10 of the collected specimens tested positive for SARS-CoV-2 infection. All passengers were thus ordered to remain in their cabins for 14 days, beginning on 5 February. Key features of this quarantine are listed in Box A. A total of 712 cases of novel coronavirus disease (COVID-19) were detected among passengers and crew with 13 resulting in death as of 20 April.

#### Box A: Key features of the quarantine at Yokohama for Diamond Princess, 2019

1. confining passengers to their cabins
2. allowing 60-minute daily walk on the deck while wearing masks and one-meter distance from other passengers, monitored by staff under the guidance of health officials
3. reducing crew services (e.g. food delivered to passengers doors, cabin cleaning suspended, linens and towels delivered to cabin doors)
4. modified infection-prevention-and-control contingency plans among crew that are typically used for norovirus
5. turning off air re-circulation and increasing extraction in cabins to prevent possible airborne transmission (cabins aboard cruise ships, independent of suspected outbreaks, maintain negative pressure)
6. postponing disinfection of affected cabins until all guests and crew had disembarked the vessel.
7. Cases were transferred from their cabins to isolation facilities. Their contacts were then tested and remained in the cabin unless a specimen obtained from them resulted RNA detection, at which point they were also transferred to a healthcare facility.

The objective of this study was to test the hypothesis that environmental surfaces, wastewater, and air played important roles in transmission for SARS-CoV-2 during this outbreak. Such information could inform outbreak prevention and control strategies as well as disinfection procedures.

## Method

This was a cross-sectional study to test environmental samples in the cruise ship. On 22 and 23 February 2020, prior to disinfection of the vessel and some passengers and crew remained aboard, we obtained specimens from cabins and common areas following the procedures outlined by the US Centers for Disease Control and Prevention for the detection of human norovirus on surfaces^5^.

Cases were defined as any person aboard the vessel from 3 to 25 February who had at least one oropharyngeal specimen test positive for SARS-CoV-2 by real-time reverse transcriptase polymerase chain reaction (rRT-PCR), independent of symptom presentation. Cases were further defined as symptomatic or asymptomatic based on their presentation at the time of respiratory specimen collection. For case-cabins, we randomly selected cabins in which confirmed symptomatic or asymptomatic COVID-19 cases had stayed. To understand the duration and survivability of SARS-CoV-2 on surfaces, we also selected case-cabins according to the last date on which any person was in the cabin. Case-cabins had been disinfected by 5% hydrogen peroxide spraying prior to sampling (February 14-15), some of which were also sampled. To understand the contribution of air transmission, we selected non-case-cabins (those with no confirmed case at any point) next to a case-cabin or at least 3 cabins away from a case-cabin. To understand the contribution of wastewater, we selected a non-case-cabin below a case-cabin. The cabins sampled included both cabins with and without windows (i.e. interior cabins). For each selected case-cabin, we swabbed these locations: the cabin light switch, doorknob, toilet flush button, toilet seat, bathroom floor, chair armrests, television remote control, telephone, desk, and bed pillow. Locations of sample sites are listed in Box B.

#### Box B: Locations swabbed

Common areas

- Handrails
- Phones
- Armrests
- Computer keyboards
- Staff elevator handrails and buttons
- Meal cart handles
- Cafeteria armrests and desks
- Doorknob of a laundry room
- Hood in the corridor

Japanese medical response headquarters in the deck 5 restaurants

- Tables
- Computer mouse
- Keyboards
- Printers
- Phones
- Coffee pots
- Milk pots
- Pens
- Handrails
- Trash box - Sofa

Onboard medical centre

- Telephone handset
- Intercom button in the lobby at the entrance
- Waiting-room chair armrest
- Waiting-room bathroom doorknob
- Interior and exterior clinic doorknobs
- Vital-sign monitors
- Oxygen flowmeters
- Portable ventilator
- Examination room chair
- Bed wall
- Examination room computer keyboard
- Portable oxygen saturation meter mounting section
- Examination room trashcan
- Computer keyboard outside examination room
- Wagon and goggles on the wagon
- Treatment room doorknob
- Blood count meter
- Alcohol disinfection equipment

We used polyester-flocked oropharyngeal specimen-collection swabs and moistened them with Viral Transport Medium (VTM). We then swabbed areas (4×5 cm^2^) in 3 directions. We placed swabs into VTM and kept them refrigerated at −80°C until submission to National Institute of Infectious Diseases, Japan (NIID). In addition, a second sampling of surfaces from part of the SARS-CoV-2 RNA-detected items was conducted for viral isolation on February 27, with the samples stored at 4°C and transferred directly to the laboratory for isolation to account for any loss to sample quality during the freezing process.

For air sampling, we selected case- and non-case-cabins. We obtained air samples from these cabins by placing two air samplers (Airport MD8, Sartorius, 50L/min for 20 minutes) in each selected cabin, on the bed and on the toilet seat, about 1.5 meters away from the handle side of the sliding door while keeping the door closed. Collection was performed through a special gelatin filter (type 175, Sartorius, T1 phage capture rate: 99.99%, effective filtration area: 38.5 cm^2^). After collection, the sample was put in the gelatin filter in the original package, checked, and stored at −80 °C until it could be transferred to NIID (typically at least 14 days).

Samples were tested by rRT-PCR according to the protocol described by NIID^6^. We then attempted viral isolation from some samples from which viral RNA was detected by rRT-PCR and second samples.

Specimens were mixed with Dulbecco’s modified Eagle medium supplemented with typical concentrations of penicillin G, streptmycin, gentamicin, amphotericin B and 5% fetal bovine serum. They were inoculated on confluent VeroE6/TMPRSS2 cells as described previously^7^. Culture medium at 0- or 48-hours post-infection (hpi) were collected and diluted 10-fold in water, then boiled for 5 minutes. A rRT-PCR assay was performed to quantify the increased amount of coronavirus RNA with a MyGo Pro instrument (IT-IS Life Science, Ireland) using primers and probes described previously^8^.

The median highest and lowest temperature in Yokohama between 3 February and 27 February were 13.0°C (range 6.5-18.5) and 5.5°C (0.0-9.3). The median highest and lowest humidity were 73 (41-98) % and 40 (17-76) %^9^.

We described the results and used Fisher’s exact test to evaluate the difference of proportion of SARS-CoV-2 detection in the cabins with symptomatic and asymptomatic cases. We considered two-tailed p < 0.05 statistically significant, and used Bonferroni correction. We used Stata 14 (Stata Corp., College Station, TX, USA) for calculation. This report was exempt from the requirement for institutional ethics review since it was a public health investigation by the Japanese Infectious Disease Law and Quarantine Law.

## Results

In total, 601 environmental samples were collected and tested, of which SARS-CoV-2 RNA was detected from 58 samples (10%) (Table 1). SARS-CoV-2 RNA was detected from approximately two-thirds of all case-cabins swabbed, while it was not detected from any of the non-case cabins. Except for one sample from an air hood in a corridor, SARS-CoV-2 RNA was not detected from samples swabbed in common areas. SARS-CoV-2 RNA was not detected from all the air sampling.

**Table 1.**
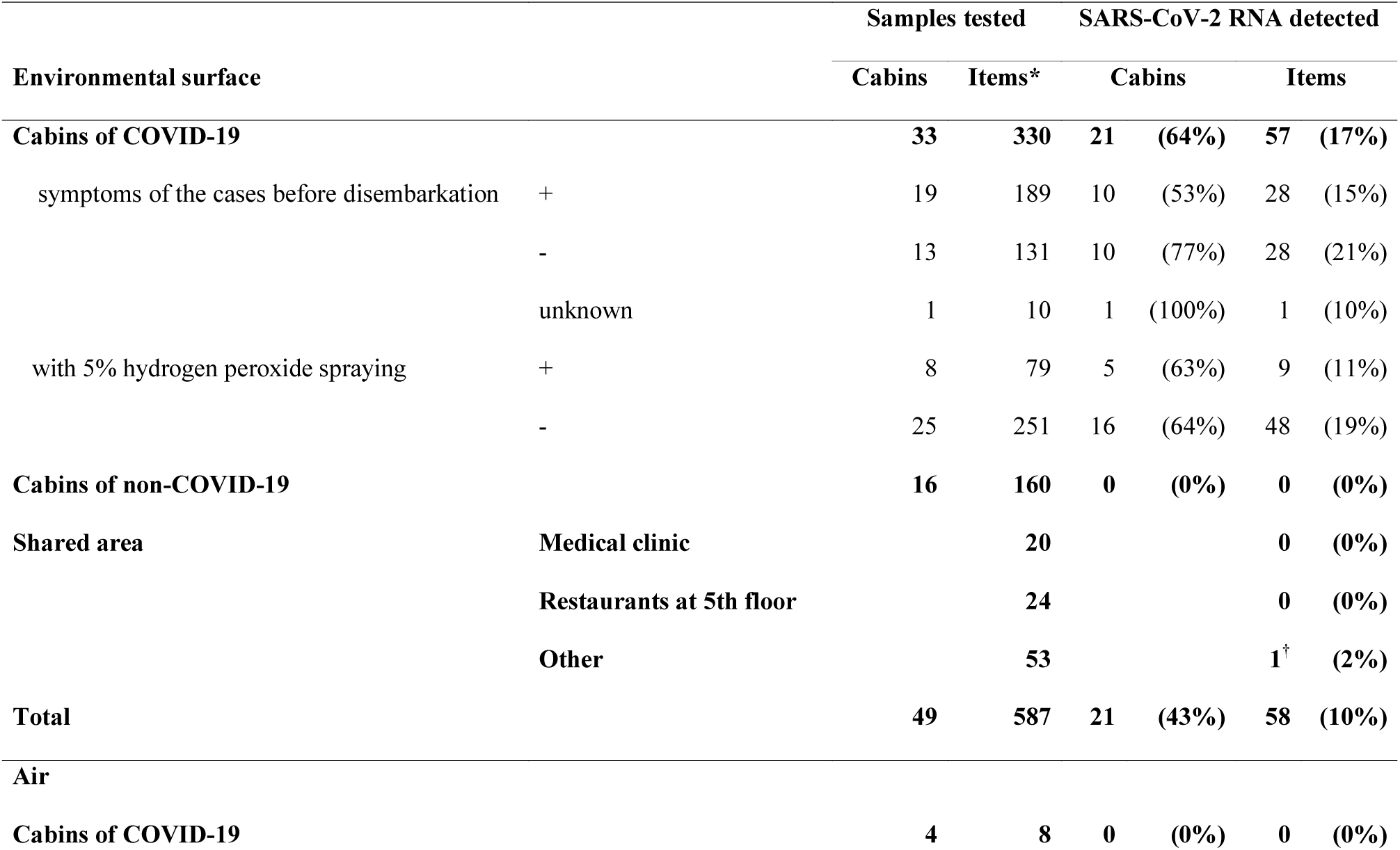
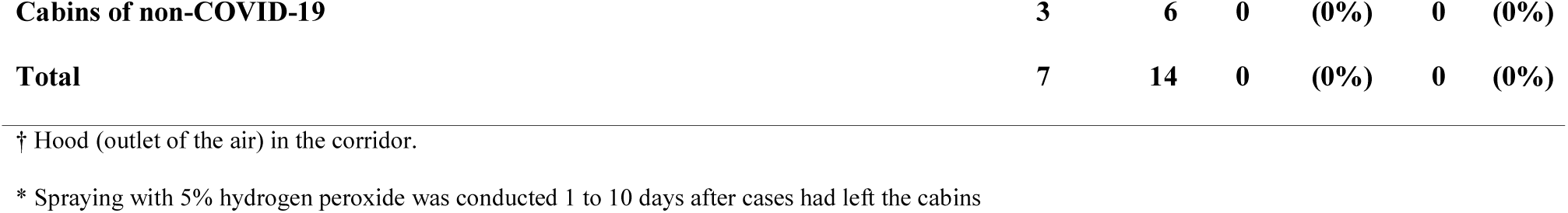
Detection of SARS-CoV-2 RNA by cabin and area.

|  |  |  | Samples tested |  | SARS-CoV-2 RNA detected |  |  |
| --- | --- | --- | --- | --- | --- | --- | --- |
| Environmental surface |  |  | Cabins | Items* | Cabins |  | Items |
| Cabins of COVID-19 |  |  | 33 | 330 | 21 | (64%) | 57 (17%) |
| symptoms of the cases before disembarkation | + |  | 19 | 189 | 10 | (53%) | 28 (15%) |
|  | - |  | 13 | 131 | 10 | (77%) | 28 (21%) |
|  | unknown |  | 1 | 10 | 1 | (100%) | 1 (10%) |
| with 5% hydrogen peroxide spraying | + |  | 8 | 79 | 5 | (63%) | 9 (11%) |
|  | - |  | 25 | 251 | 16 | (64%) | 48 (19%) |
| Cabins of non-COVID-19 |  |  | 16 | 160 | 0 | (0%) | 0 (0%) |
| Shared area | Medical clinic |  |  | 20 |  |  | 0 (0%) |
|  | Restaurants at 5th floor |  |  | 24 |  |  | 0 (0%) |
|  | Other |  |  | 53 |  |  | 1<sup>†</sup> (2%) |
| Total |  |  | 49 | 587 | 21 | (43%) | 58 (10%) |
| Air |  |  |  |  |  |  |  |
| Cabins of COVID-19 |  |  | 4 | 8 | 0 | (0%) | 0 (0%) |

|  |  |  |  |  |  |  |
| --- | --- | --- | --- | --- | --- | --- |
| **Cabins of non-COVID-19** | **3** | **6** | **0** | **(0%)** | **0** | **(0%)** |
| **Total** | **7** | **14** | **0** | **(0%)** | **0** | **(0%)** |

† Hood (outlet of the air) in the corridor.
\* Spraying with 5% hydrogen peroxide was conducted 1 to 10 days after cases had left the cabins

Table 2 presents the items from which SARS-CoV-2 RNA was detected in case-cabins. RNA was most often detected on the floor around toilet in the bathroom (39%, 13/33, cycle quantification (Cq): 26.21-37.62) and the bed pillow (34%, 11/32, Cq: 34.61-38.99).

**Table 2.**
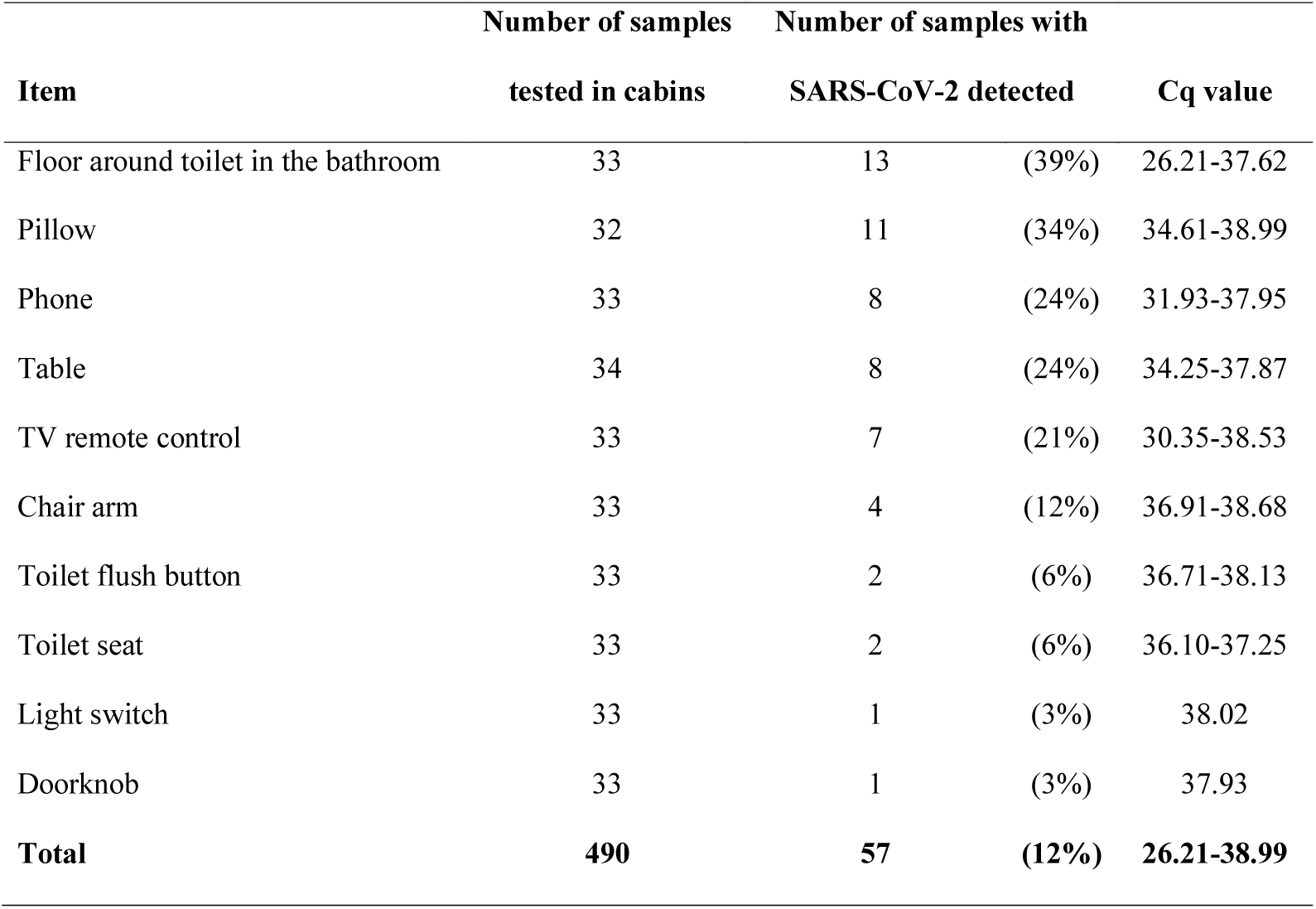
Detection of SARS-CoV-2 RNA in case-cabins by swabbed item.

In case-cabins with symptomatic persons (including symptomatic-only and mixed symptomatic/asymptomatic cabins), SARS-CoV-2 RNA was detected from 15% (28/189) of the non-case-cabins with Cq values ranging 29.79-38.86 (Table 3). In case-cabins in which only asymptomatic cases had stayed, SARS-CoV-2 RNA was detected from 21% (28/131) of the case-cabins with a range of Cq values of 26.21-38.99. All but two case-cabins had two persons staying in the room before vacating. The remaining two cabins had one or three persons stayed before vacating.

**Table 3.**
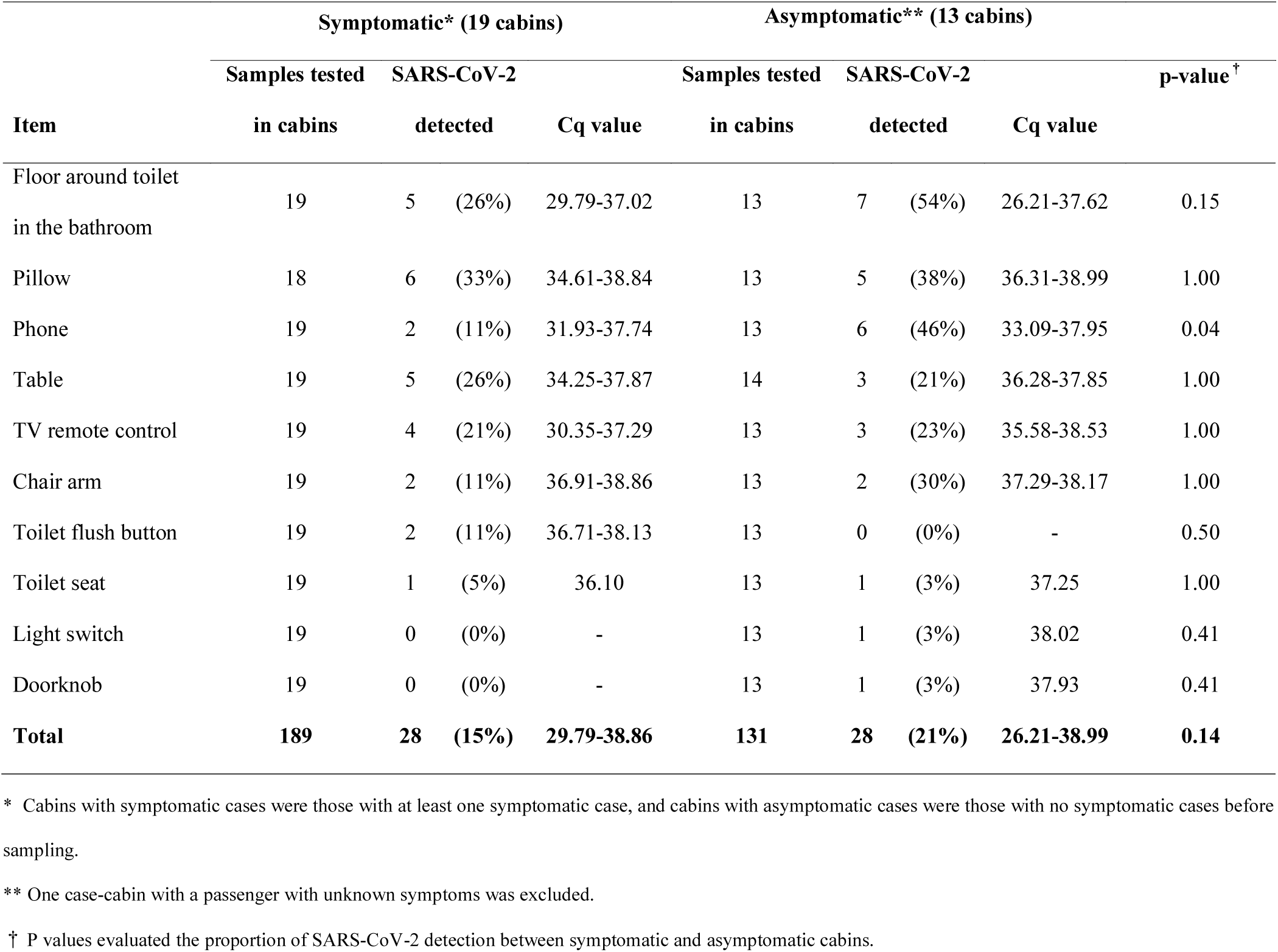
Detection of SARS-CoV-2 RNA in case-cabins by swabbed item by symptoms of the cases.

Table 4 presents the time between the last person vacating the case-cabin and the swabbing of areas. The range was 1-17 days for detection of SARS-CoV-2 RNA. Those areas that SARS-CoV-2 RNA was detected at least 14 days after the cabin was vacated were the floor around toilet in the bathroom and the pillow. The lowest Cq values were detected on samples taken four (26.21) and seven (29.79) days after the cabins were vacated. Both samples were obtained from the floor around the toilet in the bathroom.

**Table 4.**
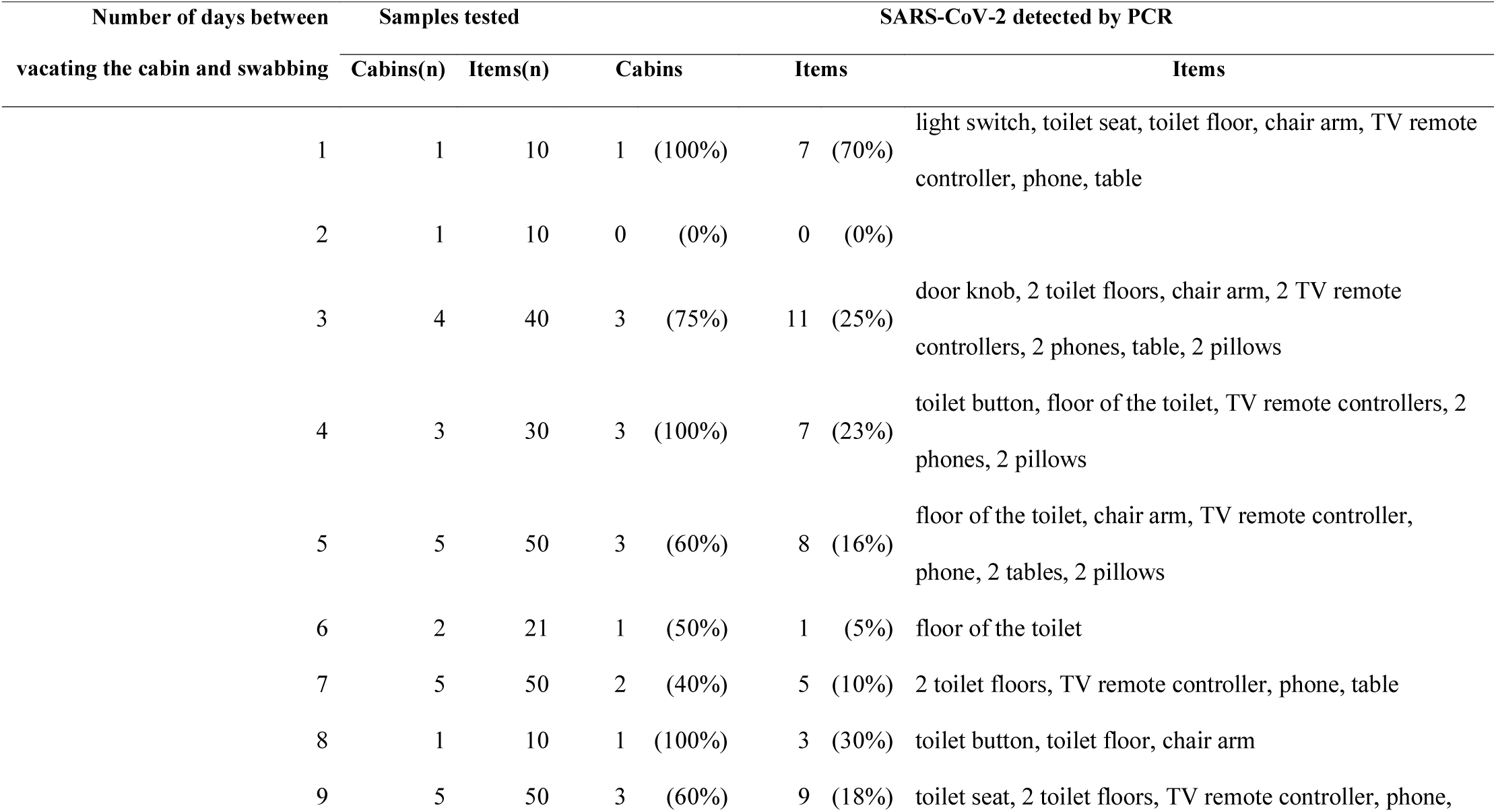
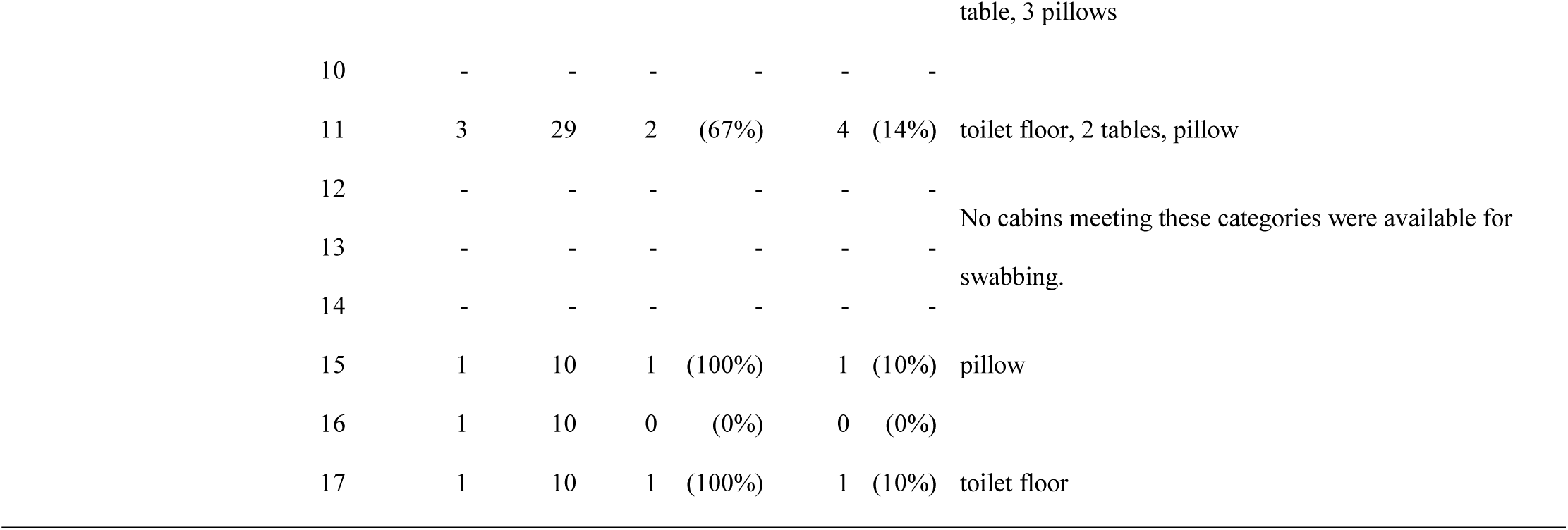
Duration of SARS-CoV-2 RNA detection on surfaces.

Among the 58 samples with SARS-CoV-2 RNA detected by rRT-PCR, none could be isolated. Among the eighteen samples obtained in the second sampling, none could be isolated.

## Discussion

After a COVID-19 outbreak that involved in 712 cases out of 3,711 persons aboard a commercial cruise vessel, we detected SARS-CoV-2 RNA on the environmental surfaces of cabins of symptomatic and asymptomatic COVID-19 cases up to 17 days after the cabins had been vacated. We did not detect SARS-CoV-2 RNA on surfaces of non-case-cabins nor on surfaces of common areas except one. Neither did we detect viral RNA in the air. Although we were unable to isolate the virus from any of the samples with SARS-CoV-2 RNA by rRT-PCR or the second samples, our findings have implications for outbreak prevention and control strategies as well as disinfection procedures.

Our findings suggest that air transmission and wastewater transmission do not play a major role in outbreaks of COVID-19. A recent air-sampling study of three COVID-19 patient rooms in a hospital found no positive air samples^10^, but another study reported that the virus are liable for up to three hours in the air^11^. The lone detection of SARS-CoV-2 RNA from an air vent in the ceiling of a corridor is more likely the result of a projectile droplet or of a hand touching the vent. Some respiratory pathogens, such as influenza virus or SARS-CoV, have been reported to transmit beyond one meter in some circumstances^12^. Alternatively, stopping the air re-circulation aboard the ship may have prevented airborne transmission in the common area or between the cabins. The effect of stopping the air recirculation in the cruise ship during COVID-19 outbreak needs further study.

Our findings suggest rather that environmental surfaces may play a role in transmission of the virus. SARS-CoV-2 RNA was detected on multiple surfaces of case-cabins, most often the pillow and the bathroom floor for up to 17 days, which was longer than previously reported^13^. In humans, SARS-CoV-2 has been detected in oral swabs, anal swabs and blood^14^, as well as tears, conjunctivae, and sputum.^15^ A recent investigation into the biodistribution of SARS-CoV-2 among 1070 COVID-19 patients showed high RNA detection rates by rRT-PCR in bronchoalveolar lavage fluid, sputum, and nasal swabs, with lower detection rates in pharyngeal swabs, feces, and blood, but no detection in urine^16^. The SARS-CoV-2 RNA detected on bed pillows of case-cabins in this cruise vessel may have come from coughing, nasal drainage, or tears during sleep. This suggests that appropriate cleaning of linens is also important for the outbreak control. The RNA detected on the floor of toilet from the bathroom may have come stool^14^ or from respiratory tract. That other surfaces with high frequency of hand-touching (e.g. doorknobs) resulted RNA detection less often may be due to good hand hygiene practices, frequent cleaning of these surfaces, or the material of which the surface was made^11^. As with health-care settings, where patient hand-hygiene guidance is essential to prevent healthcare-associated infections^17^, communication on good hand hygiene is critical for stopping transmission on cruise vessels under quarantine.

Another important implication of our findings is that cases who are symptomatic and asymptomatic at the time of specimen collection could be shedding SARS-CoV-2.^18^. As Rothe et al reported^18^, these “asymptomatic cases” may have become symptomatic or may have been post-symptomatic with barely recognizable symptoms. Nevertheless, the fact that they were asymptomatic at the time of their vacating the room implies that persons we classify as asymptomatic may be shedding. An investigation of a two-family cluster in Zhejiang Province, China, identified a potentially pre-symptomatic person—later laboratory-confirmed COVID-19—as a source of infection^19^. Asymptomatic transmission presents a substantial challenge for public health because isolation of symptomatic patients only will not interrupt the chain of transmission. In an analysis of 133 COVID-19 patients in Beijing, the authors concluded that person-to-person transmission was the main route and that controlling mild and asymptomatic cases was important for prevention^20^.

Our findings also imply that simple cleaning procedures of the environment can remove the virus from surfaces and reduce transmission. In addition to the low proportion of RNA detection on the surface samples mentioned above (e.g. door knobs), RNA was detected from only one sample in the common areas. That sample was obtained from a ceiling vent, which may have been difficult to reach during cleaning. For cleaning during the quarantine, standard disinfectant with hydrogen peroxide as the active ingredient was used, and the frequency of disinfection was increased, with a focus on areas of highest foot traffic (personal communication, J. Leonard, 19 March 2020). Although it is possible that people were in common areas when the virus was present on surfaces and could have become infected by touching those surfaces only to have those areas cleaned before we could swab them, the lack of detection of RNA in these areas reduces the relative probability of their having been virus at the time passengers and crew were there. Thus, the contribution of environmental surfaces in transmission might be limited with periodic cleaning using hydrogen peroxide products. Interestingly, SARS-CoV-2 RNA was detected in case-cabins that had been disinfected by hypochlorite spraying. Although the spraying of hydrogen peroxide could structurally disinfect SARS-C0V-2^21^, removing the virus by wiping environmental surfaces may be safer during the outbreak.

A major question that remains to be answered globally is thus the duration of viable viruses on environmental surfaces. A review of evidence on the persistence of all known coronaviruses concluded that human coronaviruses can persist on hard surfaces at room temperature for up to nine days, it did not include SARS-CoV-2^13^. A recent study indicates that SARS-COV-2 has varying viability on different surfaces and was similar to SARS-COV-1 under experimental conditions, with the virus surviving on plastics and stainless steel for over 72 hours^11^. The low Ct values in most of the detected samples suggests low level contamination of the environment after the COVID-19 cases left the place, and the low viral load in the environment may be the reason why no virus was isolated from the samples. Alternatively, there may have been some aspect of the sampling, storage, transport, or isolation method that complicated isolation success.

The strength of our study is that we took environmental sampling systematically even during in the middle of outbreak responses. Also, the rooms left untouched for days after the embarkation of passengers, which provided an ideal situation to evaluate the persistence of viral RNA. Our findings and interpretations should take into consideration the following limitations. First, it took approximately three hours to bring the specimen to the laboratory due to logistical challenge in a cruise ship, which may affect the viral isolation. Second, we could not directly measure the temperature and humidity in the cruise ship.

In conclusion, the environment around the COVID-19 cases was extensively contaminated from SARS-CoV-2 during COVID-19 outbreak in the cruise ship. The environmental surface could involve viral transmission through direct contact, but may not be through air or wastewater mechanisms. This transmission can occur from persons who are asymptomatic at the time of specimen collection. Cleaning of surfaces with hydrogen peroxide-based products and communication messages demonstrating and emphasizing hand hygiene are essential to interrupting the chain of transmission during outbreaks.

## Data Availability

All the data referred to in the manuscript are available.

## ACKNOWLEDGEMENT

We acknowledge Ministry of Health, Labour and Welfare including quarantine officers and other public health officers who helped to take samples from the environment. We thank all the crew in the Princess Cruises for their dedicated cooperation against the response. We also thank all the staff of National Institute of Infectious Diseases who worked for the COVID-19 response.

## AUTHOR CONTRIBUTION

TY, GM, MP and BB designed the study, TY, KW, RF NO, TG, NM, MS, TT, ST, HS, KO sampled from the environment, TW, MO, KO, NN, KS, SM, TM, MT, TK, HH conducted laboratory testing of the samples, TY, MO, GM, PA, BB analyzed the data and developed a draft manuscript, NO, TW, MO, KH, KK, HK, AS, MS, TK, RF, JL reviewed the manuscript and provided inputs.

